# Immune transcriptomes of highly exposed SARS-CoV-2 asymptomatic seropositive versus seronegative individuals from the Ischgl community

**DOI:** 10.1101/2020.09.01.20185884

**Authors:** Hye Kyung Lee, Ludwig Knabl, Lisa Pipperger, Andre Volland, Priscilla A. Furth, Keunsoo Kang, Harold E. Smith, Knabl Ludwig, Romuald Bellmann, Christina Bernhard, Norbert Kaiser, Hannes Gänzer, Mathias Ströhle, Andreas Walser, Dorothee von Laer, Lothar Hennighausen

## Abstract

To investigate prevalence of ongoing activation of inflammation following asymptomatic SARS-CoV-2 infection we characterized immune cell transcriptomes from 43 asymptomatic seropositive and 52 highly exposed seronegative individuals with few underlying health issues following a community superspreading event. Four mildly symptomatic seropositive individuals examined three weeks after infection as positive controls demonstrated immunological activation. Approximately four to six weeks following the event, the two asymptomatic groups showed no significant differences. Two seropositive patients with underlying genetic disease impacting immunological activation were included (Cystic Fibrosis (CF), Nuclear factor-kappa B Essential Modulator (NEMO) deficiency). CF, but not NEMO, associated with significant immune transcriptome differences including some associated with severe SARS-CoV-2 infection (IL1B, IL17A, respective receptors). All subjects remained in their usual state of health from event through five-month follow-up. Here, asymptomatic infection resolved without evidence of prolonged immunological activation. Inclusion of subjects with underlying genetic disease illustrated the pathophysiological importance of context on impact of immunological response.

## Background

Coronavirus disease 2019 (COVID-19) is caused by severe acute respiratory syndrome coronavirus (SARS-CoV) 2. Clinical presentation ranges from asymptomatic to severe disease. In symptomatic patients, increased levels of pro-inflammatory cytokines and subsequently lymphopenia are reported coincident with disease^1,2^. Flow cytometry-based^3^ and single-cell transcriptome studies^4-7^ defined inflammatory milieus with an overt innate and adaptive immune activation and immune signatures of COVID-19 patients with different disease trajectories^7-10^.

On occasion COVID-19 patients can suffer from longer term sequelae. In one survey, thirty-five percent of patients with mild outpatient-treated disease were reported as having not returned to their usual state of health by two to three weeks following infection^11^. Prolonged myocardial inflammation^12^ and subacute thyroiditis^13^ post resolution of acute infection have been reported. Although it is increasingly being recognized that up to 96% of infected individuals are asymptomatic^14,15^, their immune responses and the prevalence of unrecognized ongoing inflammation have not been investigated and are not understood.

With the pandemic extending itself into populations with underlying genetic disease, there is an urgent need to understand their immune response to SARS-CoV-2 infections. Towards this end we determined the immune transcriptome of an asymptomatic seropositive patient with Cystic Fibrosis (CF) and one with Nuclear factor-kappa B Essential Modulator (NEMO) deficiency within our study population.

## Results

In an attempt to define the immune response in asymptomatic SARS-CoV-2 seropositive and highly exposed seronegative individuals within an isolated population, we conducted a study on residents from the ski resort of Ischgl that experienced a superspreading event in early March of 2020. This explosive local outbreak led to the spread of the virus throughout Austria and to many other European countries and worldwide^16^. Ischgl and the Paznaun valley were quarantined on March 13, 2020 and remained under lockdown for six weeks. An epidemiologic study targeting 79% (n=1473) of the population of Ischgl (n= 1867) was conducted between April 21 and 27 and revealed a seroprevalence of approximately 42% (Knabl et al., High SARS-CoV-2 Seroprevalence in Children and Adults in the Austrian Ski Resort Ischgl, *submitted)* with approximately 17% of these being asymptomatic. Our study encompassed 43 seropositive asymptomatic individuals (Group A) and 52 highly exposed seronegative individuals (Group B) with an equal gender and age distribution (Fig. 1a and Table 1). Six households had both seropositive asymptomatic and highly exposed seronegative members (Supplementary Table 1). Only a few asymptomatic seropositive individuals had conditions that increased their risk of severe illness from COVID-19^17^, one patient with Cystic Fibrosis *(CFTR* G551D mutation) and one with Nuclear factor-kappa B Essential Modulator (NEMO) deficiency (Incontinentia pigmenti, *IKBKG* exon4_10del mutation) (Table 1). All infected individuals in the study remained asymptomatic throughout their infection and were confirmed in in person interviews as having their usual state of health at the time of the study (4-6 weeks after the infection) by telephone and remained so at the second phone call five months following the outbreak.

**Fig. 1.**
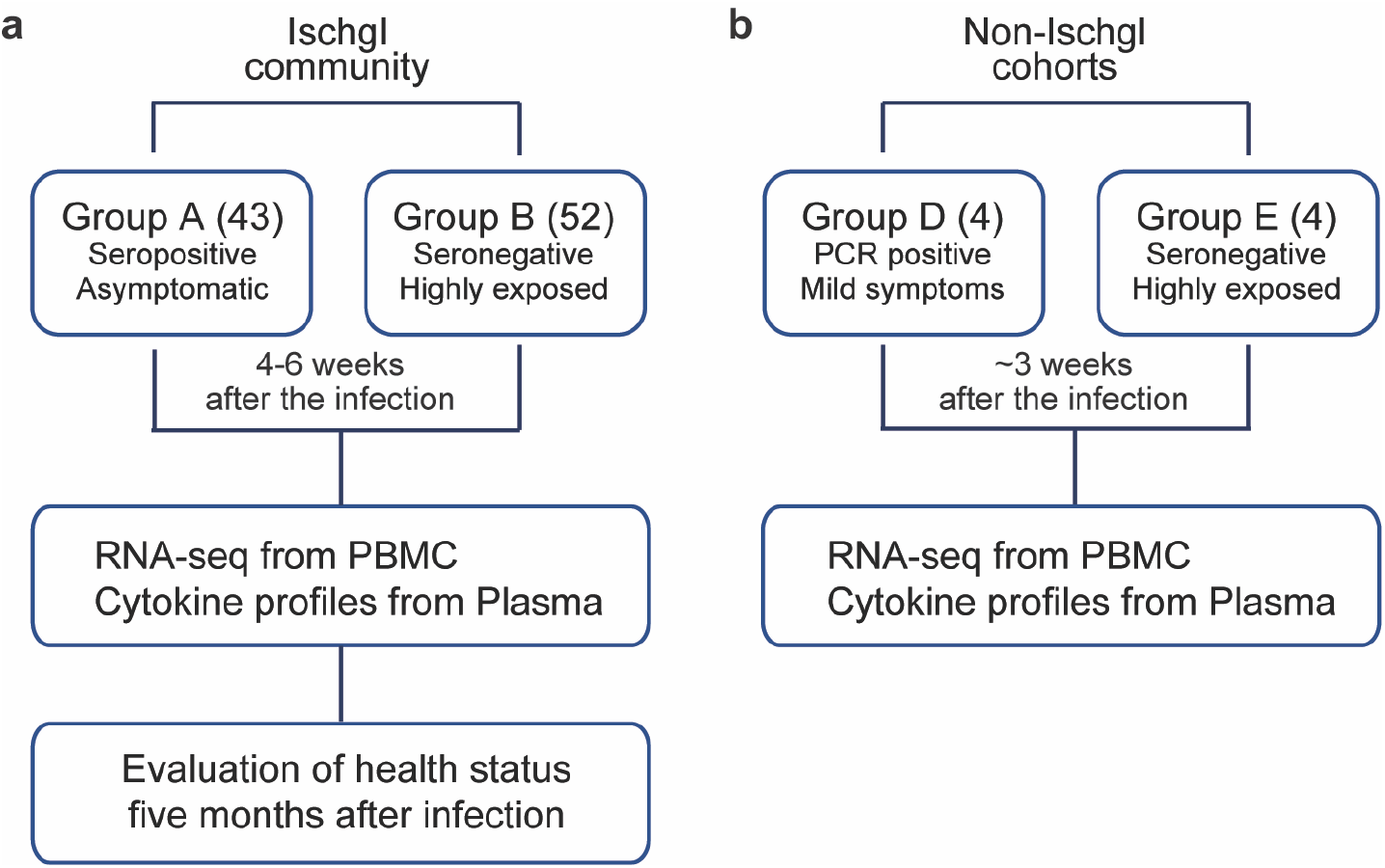
Study Design. **a** In the Ischgl community, 43 seropositive asymptomatic and 52 highly exposed seronegative individuals underwent phlebotomy and health evaluation between April 21 and 27, 2020 with a follow up health evaluation mid-August, 2020. The SARS-CoV-2 superspreading event occurred in the community between the end of February and March 13 of 2020 when the town was quarantined. **b** The non-Ischgl cohorts were recruited from other parts of Tyrol. Four SARS-CoV-2 PCR positive patients exhibiting mild symptoms who underwent phlebotomy and health evaluation approximately three weeks after being confirmed as PCR positive for SARS-CoV-2 and four highly exposed seronegative individuals, from the same household as one of the symptomatic patients were included.

To validate that RNA sequencing (RNA-seq) performed on RNA extracted from peripheral blood mononuclear cells (PBMCs) could be used to identify gene expression changes in patients following SARS-CoV-2 infection, we conducted an unbiased RNA-seq analysis on PBMCs from four patients with mild symptoms (Group D), about three weeks after they had tested PCR-positive. These were compared to four highly exposed seronegative individuals (Group E) (Fig. 1b and Supplementary Table 2), who cohabitated with one of the symptomatic patients. Among the 175 genes whose expression was significantly elevated in PBMCs from symptomatic patients, genes were significantly enriched in 16 Hallmark gene sets (Supplementary Table 2), five of which have defined roles in immune regulation: TNF-α/NFκB, mTORCI signaling, IL2-STAT5 signaling, TGFβ signaling and inflammatory response (Fig. 2 and Supplementary Table 2). This demonstrates that the RNA-seq approach utilizing buffy-coat-isolated PBMCs identified statistically significant differences in inflammatory gene expression between infected and non-infected individuals. Notably, we observed significantly elevated expression of IL10 in the symptomatic patients, a cytokine whose elevated expression has been associated with disease severity^18^.

**Fig. 2.**
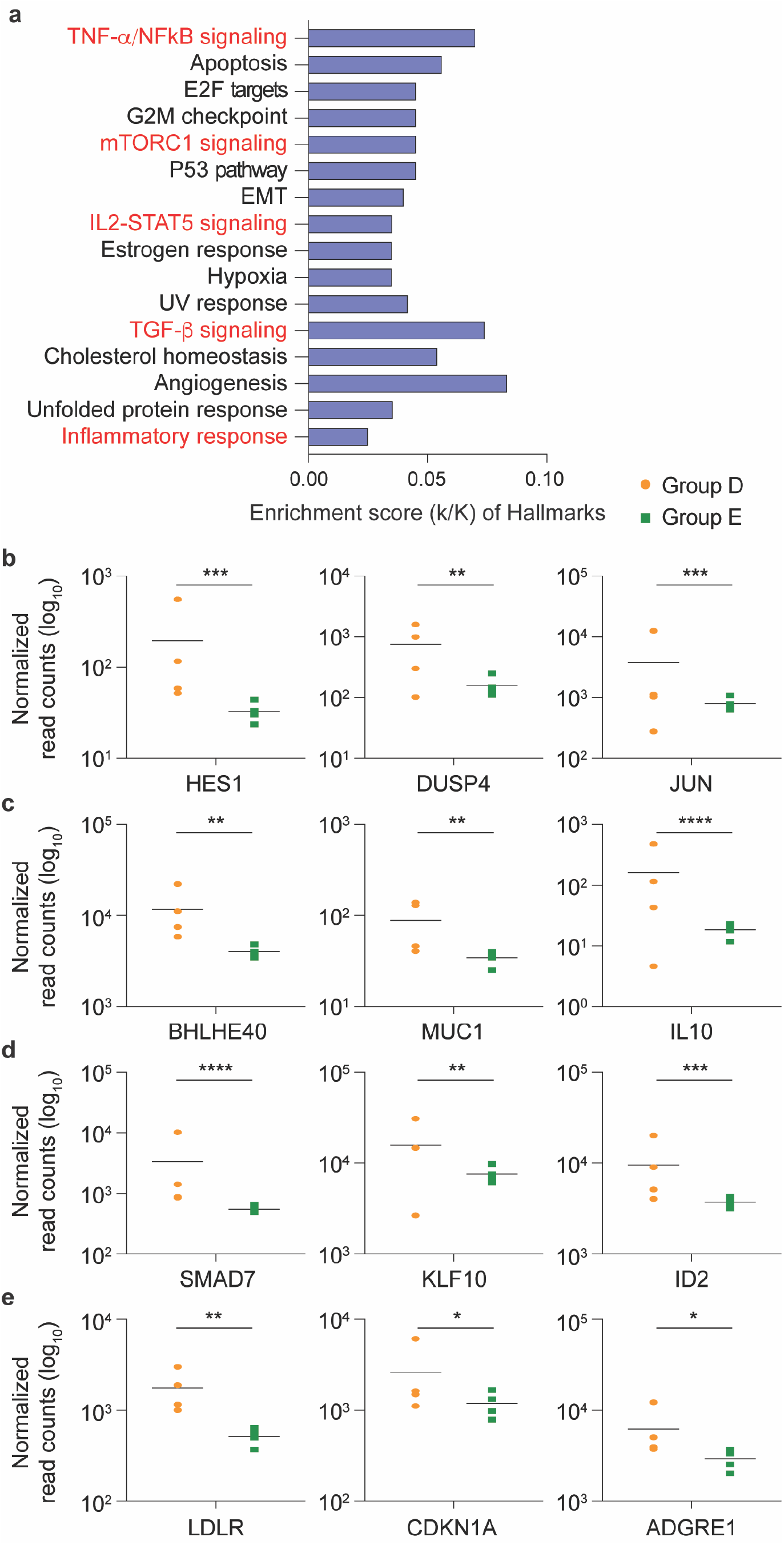
SARS-CoV-2 infected patients with mild symptoms demonstrated significant elevations in immune response genes three weeks following PCR confirmation of SARS-CoV-2. **a** Genes expressed at significantly higher levels in the infected patients were significantly enriched in 16 Hallmark Gene Sets (FDR q-value < 0.006). Five are involved in immune regulation: TNF-α NFκB, mTORC1, IL2-STAT5, TGF-β, inflammatory response (labeled in red). The other 11 gene sets are not directly linked to immune response. **b-e** Comparison of relative normalized gene expression levels of three representative genes from each of the four immune regulation-related Hallmark Gene Sets between infected (orange dots, Group D, n=4) and non-infected (green dots, Group E, n=4) individuals. Mean indicated. \**P*adj < 0.04, \*\**P*adj < 0.001, \*\*\**P*adj < 0.0001, \*\*\*\**P*adj < 0.00001 (DESeq2).

We then conducted an unbiased RNA-seq transcriptome analysis (Supplementary Table 3) and plasma cytokine profiling (Fig. 3 and Supplementary Table 4) from the asymptomatic seropositive (Group A) and highly exposed seronegative (Group B) Ischgl resident cohorts. Very few statistically significant changes in gene expression were found (11 induced, 7 down-regulated) (Supplementary Table 3). Quantification of inflammatory cytokines and chemokines revealed no significant differences in protein levels between Group A and B (Fig. 3 and Supplementary Table 4).

**Fig. 3.**
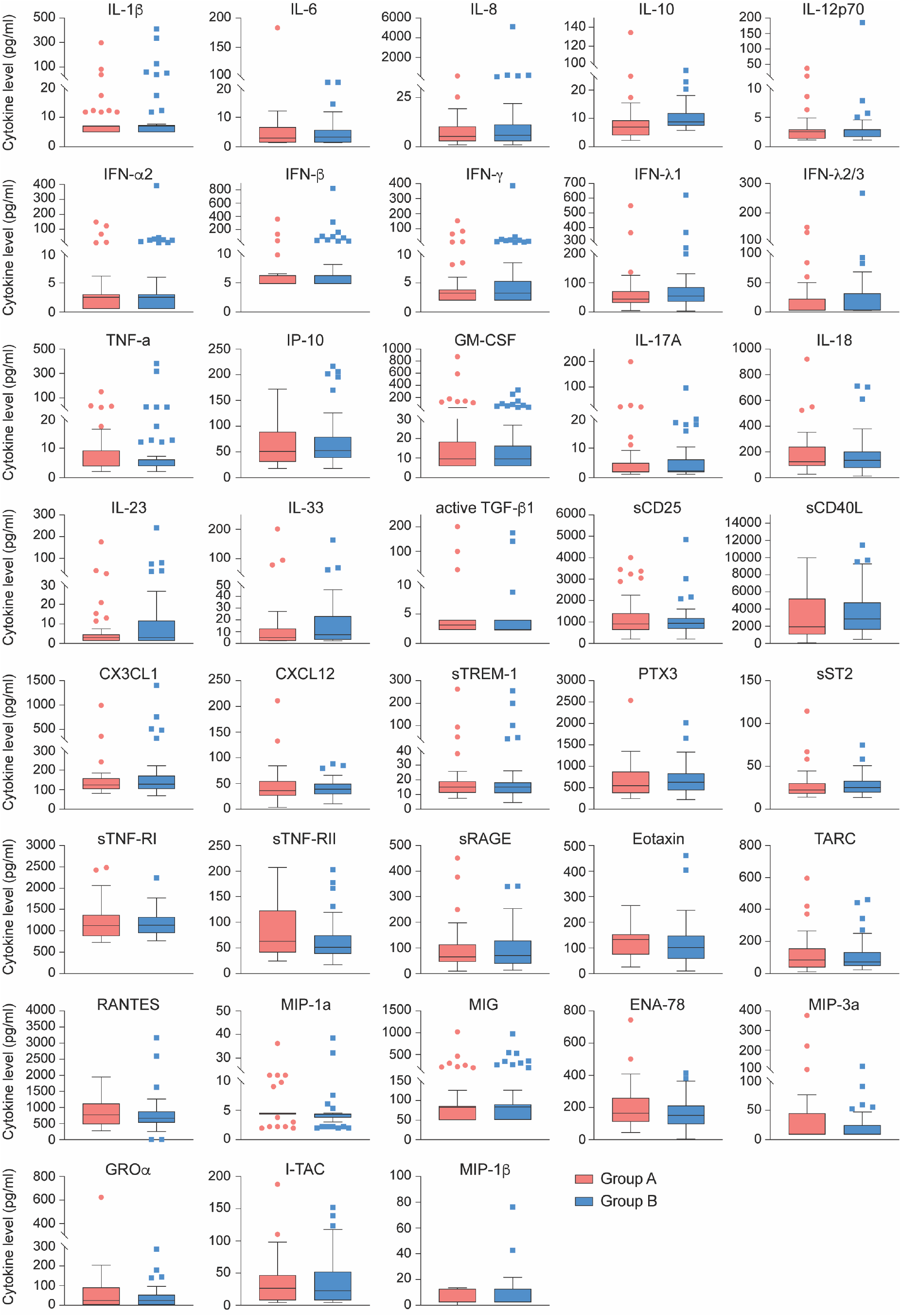
Comparison of cytokine and chemokine levels between asymptomatic seropositive and highly exposed seronegative cohorts. No statistically significant differences in relative steady state protein levels of 28 cytokines and 10 chemokines between asymptomatic seropositive (red, Group A, n=41) and highly exposed seronegative (blue, Group B, n=52) were found. Relative protein levels measured using LEGENDplex assays. Boxplots show median (middle bar), interquartile range (IQR) (box), 1.5 X IQR (whiskers).

The PBMC transcriptome data from the CF (Supplementary Table 5) and NEMO (Supplementary Table 6) patients in Group A were analyzed separately. While the NEMO patient showed no statistically significant differences as compared to Group A, the CF patient demonstrated statistically significant differences in expression for approximately 4670 genes in comparison with Group A. Overall, expression levels of 3020 genes were significantly higher and expression levels of 1648 genes were significantly lower (Supplementary Table 5). Genes were enriched in 32 Hallmark gene sets, 11 of which have defined roles in immune regulation (Fig. 4a). Expression of key immune signature genes, including interferon response genes, IL1B, IL17A and their respective receptors, and JAK-STAT pathway genes, were significantly induced (Fig. 4b-d).

**Fig. 4.**
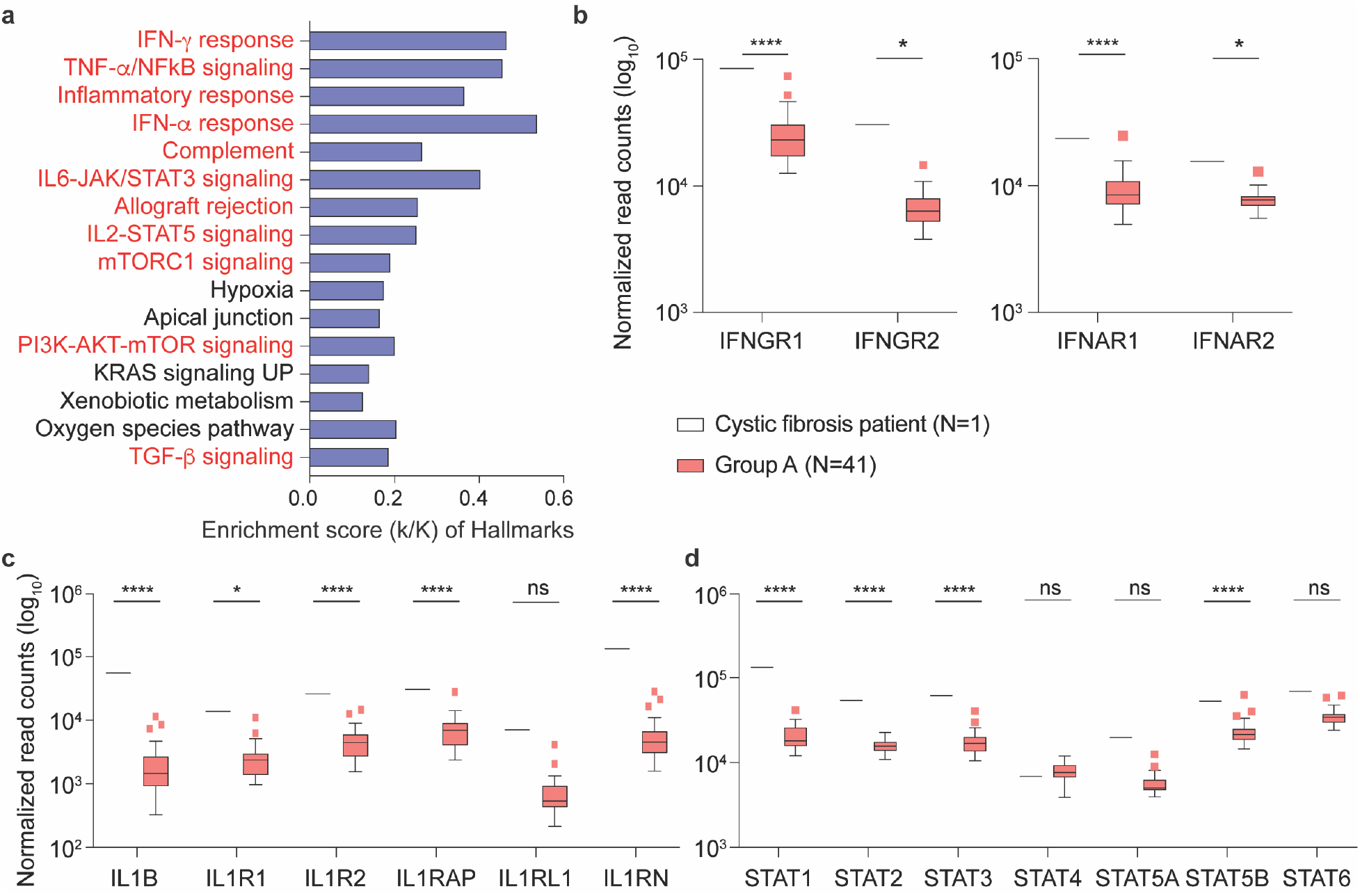
Comparison of gene expression levels between the Cystic Fibrosis patient and the remainder of the asymptomatic seropositive cohort. **a** Genes expressed at significantly higher levels in the Cystic Fibrosis patient were significantly enriched in Hallmark Gene Sets (FDR q-value < 0.005). Out of the top 16, eleven are involved in immune regulation: IFN-g, TNF-α via NFκB, inflammatory response, IFN-α, complement, IL6-JAK/STAT3, allograft rejection, IL2-STAT5, mTORC1, PI3-AKT-mTOR, TGF-β (labeled in red). The other five gene sets are not directly linked to immune response (labeled in black). **b-d** Comparison of relative normalized gene expression levels from IFN-γ (IFNGR1, IFNGR2), IFN-α (IFNAR1, IFNAR2) and TNF-α inflammatory signal (IL1B, ILR1, ILR2, IL1RAP, IL1RL1, ILRN) response and STAT family genes (STAT1, STAT2, STAT3, STAT4, STAT5A, STAT5B and STAT6). Boxplots show median (middle bar), interquartile range (IQR) (box), 1.5 X IQR (whiskers). \**P* < 0.05, *\*\*\*\*p* < 0.00001, ns, not significant.

## Discussion

The ability to assess the immune response of both symptomatic and asymptomatic SARS-CoV-2 infected individuals might provide critical information on dysregulated immune-response signatures that could foretell disease trajectories. While longitudinal studies on hospitalized patients demonstrate elevated levels of pro-inflammatory cytokines and signatures associated with ongoing inflammation^3^, there is a parallel need to pinpoint the immune response of infected, yet asymptomatic individuals. With the exception of a CF patient, we did not detect an aberrant immune transcriptome in 41 asymptomatic seropositive individuals that were infected during a super spreading event as compared to 52 highly exposed seronegative individuals from the same community. Several households housed both seropositive and seronegative individuals. In contrast to the seropositive asymptomatic individuals, a proinflammatory immune signature was detected in patients with mild symptoms. These results demonstrate that development of an antibody response to COVID-19 following viral exposure and seroconversion in asymptomatic cases is not necessarily associated with sustained alterations in the immune system transcriptome.

While other studies have focused on single cells RNA-seq (scRNA-seq) profiling of small numbers of individuals with COVID-19 disease^4-7,9,10^, the use of RNA-seq from mononuclear cells permitted us to analyze larger cohorts at a great depth. This approach was validated through the detection of inflammatory immune signatures in COVID-19 patients exhibiting mild symptoms and one asymptomatic seropositive CF patient.

Patients with CF manifest cytokine dysfunction and hyperinflammation that overlaps with the pathophysiology of COVID-19^19^. While there are limited data on the immune response of CF patients to COVID-19 infection, preliminary information suggests that the course of disease may be milder than expected^20,21^. The immune transcriptome of the asymptomatic seropositive CF patient provided evidence of highly activated cytokine signaling pathways. Expression of key components of interferon, interleukin and JAK-STAT pathways is highly elevated. Many of these genes, such as IL1B and its receptors are also highly activated in COVID-19 patients^9^. In contrast, the immune transcriptome of a SRAS-CoV-2 infected asymptomatic patient with a NEMO deficiency syndrome, a rare primary immunodeficiency, was indistinguishable from asymptomatic seropositive controls. It remains to be understood how elevated cytokine signaling, documented here in an asymptomatic CF patient, contributes to disease progression in non-CF patients and why CF patients, with chronic high expression, do not invariably experience disease progression. A critical question for development of targeted therapies is to discern between direct pathogenic immune determinants of severe disease and correlates of inflammation.

Limitations of our study include the translatability of our findings to other populations. There were few underlying health issues in this rural alpine population living at an altitude of 1,400 meters. For example, obesity, which is associated with an inflammatory state and is recognized as risk factor for severe COVID-19 disease^22,23^, was less than 10% in our study population, differing greatly from higher prevalence rate in other infected populations. Similarly, other defined risk factors for severe disease such as diabetes and chronic kidney disease, were also comparatively low.

## Methods

### Study population, study design and recruitment

The ethical committee of the Medical University of Innsbruck approved the study (EC numbers: 1100/2020 and 1111/2020), which took place between April 21 and 27, 2020. This cross-sectional epidemiological survey targeted all residents of Ischgl/Tyrol irrespective of age and gender. At the time of investigation, Ischgl had a population size of 1,867 individuals, 1,617 with their main residence in Ischgl and 250 seasonal immigrant workers, living in 582 different households.

### Quantification of immunoproteins

Samples from all groups were measured for the presence and quantity of several human inflammatory cytokines/chemokines, key targets, which are involved in inflammation and immune response and human proteins that are involved in response to viral infections. For this purpose, participants’ plasma was examined by using four different pre-defined LEGENDplex™ assays (BioLegend, California, USA), while the human inflammation panel 1 was used to quantify 13 human inflammatory cytokines including IL-17A, IL-18, IL-23 and IL-33, the pre-defined human inflammation panel 2 quantified TGF-β1, sTREM1, PTX-3, sCD40L, sCD25, CXCL12, sST2, sTNF-RI, sTNF-RII, sRAGE and CX3CL1. The human anti-virus response panel measured quantities of type 1 interferons (INF-α2, IFN-β), type 2 interferons (IFN-γ), and type 3 interferons (IFN-λ1, IFN-λ2/3) as well as the interleukins IL-1β, IL-6, IL-8, IL-10, IL-12 and TNF-α, IP-10 and GM-CSF. Additionally, chemokines were analyzed using the human proinflammatory chemokine panel. The LEGENDplex assays were used according to the manufacturer’s instructions. In short, immunoprotein capturing by antibody-bearing beads was done by using V-bottom plates. Bead-bound proteins were measured with a FACS CantoII cytometer (Becton Dickinson, New Jersey, USA), whereby fluorescent signal intensity was proportional to the amount of bead-bound proteins. The concentrations of the immunoproteins were calculated with the LEGENDplex™ data analysis software.

### Extraction of the buffy coat and purification of RNA

Extraction of the buffy coat and subsequent RNA purification will be performed as described^10^. In short, the drawn blood is centrifuged at 1,600g for 10 min at 4°C. After vacuming off the plasma layer, the buffy coat layer is carefully collected. The obtained buffy coat is mixed with 1 mL RBC lysis buffer and incubated for 10 min at room temperature. After a centrifugation step, supernatants were discarded, and the pellets mixed with 1 mL RBC lysis buffer. The pellet is washed with PBS buffer and then mixed with 1 mL TRIzol^®^. For extraction of the RNA the TRIzol reagent single-step method will be used^11^. After addition of 2 M sodium acetate (pH 4), the tube is mixed thoroughly by invertion before adding chloroform/isoamyl alcohol (49:1) and another mixing step. The sample gets incubated on ice for 15 min and subsequently centrifuged for 20 min at 10,000g and 4°C. The obtained aqueous phase is transferred to a new Eppendorf tube, 1 mL isopropanol is then added, which is followed by an incubation at −20°C for 1 h to precipitate RNA. The final RNA precipitate is won by centrifugation at 10,000g at 4°C and discarding of the supernatant. Pellets are stored at −80°C until dispatch.

### mRNA sequencing (mRNA-seq) and data analysis

Nanophotometer (Implen) was used to analyze each sample for concentration and the RNA quality was assessed by an Agilent Bioanalyzer 2100 (Agilent Technologies). The Poly-A containing mRNA is purified by poly-T oligo hybridization from 1 μg of total RNAs and cDNA was synthesized using SuperScript III (Invitrogen). Libraries for sequencing were prepared according to the manufacturer’s instructions with TruSeq Stranded mRNA Library Prep Kit (Illumina, RS-20020595) and paired-end sequencing was done with a NovaSeq 6000 instrument (Illumina).

mRNA-seq read quality control was done using Trimmomatic^24^ (version 0.36) and STAR RNA-seq^25^ (version STAR 2.5.4a) using 150bp paired-end mode was used to align the reads (hg19). HTSeq^26^ was to retrieve the raw counts and subsequently, R (https://www.R-project.org/), Bioconductor^27^ and DESeq2^28^ were used. Additionally, the RUVSeq^29^ package was applied to remove confounding factors. The data were pre-filtered keeping only those genes, which have at least ten reads in total. Genes were categorized as significantly differentially expressed with an adjusted p-value (pAdj) below 0.05 and a fold change > 2 for up-regulated genes and a fold change of < −2 for down-regulated ones. The visualization was done using dplyr (https://CRAN.R-project.org/package=dplyr) and ggplot2^30^. The genes were cutoff in the standard, less than 5 value and less than 1 log_2_ fold change and then conducted gene enrichment analysis (https://www.gsea-msigdb.org/gsea/msigdb).

### Statistical analysis

For comparison of protein levels, single sample pairs were evaluated with the unpaired two-tailed t-test to compare the distributions of two groups with the Welch’s t test to compare the two distributions (GraphPad PRISM version 8.2.0). For comparison of RNA expression levels between CF patient and asymptomatic seropositive cohort, a two-way ANOVA followed by Tukey’s multiple comparisons test was used (GraphPad PRISM). *P*<0.05 was considered statistically significant.

### Data availability

The RNA-seq data for patients will be uploaded in GEO.

## Data Availability

The RNA-seq data for patients will be uploaded in GEO.

## Acknowledgements

Our gratitude goes to the individuals from the Ischgl community who contributed to this study to advance our understanding of COVID-19. This work utilized the computational resources of the NIH HPC Biowulf cluster (http://hpc.nih.gov). NGS was conducted in the NIH Intramural Sequencing Center, NISC (https://www.nisc.nih.gov/contact.htm).

## Author contribution

Study design: LH, LK and HKL; patient recruitment and care: LK, LK Sr, RB, SB, NK, HG, MS, AW, DvL; RNA preparation: AV; cytokine assays: LP; RNA-seq analysis: HKL; Transcriptome analysis and data interpretation: HKL, LK, LH and PAF; manuscript generation; HKL, LK, PAF and LH. All authors approved the final version.

